# Baseline Severity and Early Multiorgan Failure to Predict In-Hospital Mortality in Critically Ill COVID-19 Patients: A Retrospective Cohort Study

**DOI:** 10.64898/2026.11.03.26367211

**Authors:** Ana B. Oñoro-Morales, Emilia Roy-Vallejo, Fernando Pozo Ocampo, Rocío Molina Montero, Rafael Blancas Gómez-Casero, Diego A. Rodríguez-Serrano

## Abstract

**Background:** Intensive Care Unit (ICU) mortality remains high among patients with severe coro-navirus disease. Static admission severity scores may fail to capture the dynamic clinical trajectory of critically ill patients. We hypothesized that early multiorgan failure (MOF) at 24 hours would improve mortality discrimination beyond static admission severity alone.

**Methods:** Retrospective, single-center cohort study of 345 adults with severe COVID-19 pneumonia admitted to a Spanish ICU (March 2020–December 2022). The primary outcome was all-cause in-hospital mortality. Univariate comparisons between survivors and non-survivors were performed, followed by multivariable logistic regression to identify independent predictors of mortality. Three sequential models were constructed and compared by Area Under the Receiver Operating Characteristic Curve (AUC-ROC) discrimination.

**Results:** Overall, in-hospital mortality was 20.6%. Non-survivors were older and had higher Simplified Acute Physiology Score (SAPS) II and Acute Physiology and Chronic Health Evaluation (APACHE) II scores (p<0.001). MOF at 24 hours was more frequent among non-survivors than survivors (88.4% vs. 61.7%, p<0.001). The baseline model (age, sex, Chronic Obstructive Pul-monary Disease (COPD), hypertension, and SAPS II) achieved an AUC of 0.798 (95% CI 0.742– 0.852). The final model incorporating MOF at 24 hours achieved an AUC of 0.812 (95% CI 0.754– 0.860); early MOF nearly tripled the odds of death (OR 2.78, 95% CI 1.22–6.33, p=0.015).

**Conclusions:** Combining baseline severity with early multiorgan dysfunction provides a simple, interpretable approach to mortality risk stratification in critically ill COVID-19 patients; however, external validation is needed before it can inform escalation-of-care or treatment-limitation decisions in clinical practice.

## 1. Introduction

The coronavirus disease (COVID-19) pandemic challenged Intensive Care Units (ICUs) worldwide, leading to high mortality rates among patients with critical Severe Acute Respiratory Syndrome Coronavirus-2 infection (SARS-CoV-2) [1]. However, initial estimates varied widely across individual cohorts, from 4.5% to 69.5%, with pooled figures around 32–34%[1,2].

Although overall mortality declined significantly after the first pandemic wave as ventilatory strategies, standardized pharmacological therapies, and vaccination coverage improved [3] this survival benefit was not consistently sustained across subsequent epidemic waves, with mortality rates rising again during later surges [4].

From the early stages of the pandemic, it became evident that survival in critically ill patients is strongly determined by their baseline physiological reserve and chronic vulnerability, with underlying conditions such as chronic obstructive pulmonary disease (COPD) and arterial hypertension being key prognostic factors [5,6]. Despite the marked decrease in absolute incidence as COVID-19 has settled into a more endemic pattern, its associated mortality remains high among comorbid patients, keeping severe SARS-CoV-2 infection a clinically relevant challenge that demands continued attention in the hospital and ICU setting [7].

Ever since then, ICU admission criteria have broadened, to accommodate an increasingly older and comorbid patient population, reflecting demographic aging and advances in the management of oncologic, hematologic, and other chronic conditions [8]. These changes blur traditional boundaries between hospital wards and ICU, making accurate and rapid prognostic stratification essential to guide clinical decision-making and define appropriate goals of care.

Traditionally, intensivists rely on static severity scores calculated at admission, such as Simplified Acute Physiology Score II (SAPS II) or Sequential Organ Failure Assessment score (SOFA) [9,10]. However, these fail to capture the highly dynamic clinical course of severe COVID-19, characterized by rapid endothelial injury and a dysregulated systemic inflammatory response capable of precipitating multiorgan dysfunction within hours to days of clinical worsening [11,12]. Consistent with this, the prognostic weight of individual clinical variables has been shown to shift substantially over the course of hospitalization [13].

This dynamic complexity is best captured by monitoring early clinical trajectory; specifically the development of Multiorgan Failure (MOF) within the first 24 hours, which has emerged as a stronger prognostic indicator than baseline severity alone [14,15]. Despite this, most new predictive tools incorporate complex machine-learning algorithms or hard-to-obtain laboratory biomarkers which lack bed-side utility [16,17].

We hypothesized that incorporating early multiorgan failure (MOF) at 24 hours would improve mortality discrimination beyond static admission severity alone. The objective of this study is to develop a pragmatic, bedside predictive model for in-hospital mortality in critically ill COVID-19 patients, integrating baseline severity and chronic vulnerability with early clinical trajectory. Given the increasingly older and comorbid profile of patients now requiring intensive care, such a tool could potentially help guide discussions around escalation of therapeutic effort and appropriate goals of care, in addition to its use in daily clinical practice.

## 2. Materials and Methods

### 2.1 Study Design and Setting

This retrospective observational cohort study was conducted at the Hospital Uni-versitario Príncipe de Asturias (Madrid, Spain), a 20-bed multidisciplinary ICU. The study period extended from March 2020 to December 2022, covering con-secutive waves of the COVID-19 pandemic. The study followed the **STROBE** guidelines for observational research.

### 2.2 Patient Selection

All consecutive adult patients (≥18years) admitted to the ICU with severe COVID-19 pneumonia, confirmed by a positive RT-PCR or antigenic test for SARS-CoV-2, were included. To ensure the cohort reflected the specific pathophysiology of COVID-19, patients whose primary reason for admission was unrelated to the virus (incidental infection) were excluded. Patients transferred from or to other hospitals were also excluded to guarantee a complete follow-up of the clinical trajectory within our institution.

### 2.3 Data Collection and Variables

Clinical data were retrieved from electronic medical records and the institutional ICU registry and were anonymized prior to analysis. We collected a comprehen-sive set of variables:

#### 2.3.1 Demographics and Comorbidities

Age, sex, body mass index (BMI), and comorbidities (hypertension, diabetes, obesity, COPD, cardiovascular disease, and chronic kidney disease). The Charlson Comorbidity Index was calculated to assess the overall burden of pre-existing conditions.

#### 2.3.2 Laboratory markers and microbiology parameters

Complete blood count, coagulation (platelets, D-dimer), renal and liver function tests, and inflammatory markers (C-reactive protein (CRP), procalcitonin, Lactate Dehydrogenase (LDH), and Interleukin-6) measured using the hospital’s standard clinical laboratory immunoassay as part of routine care.

SARS-CoV-2 variants were determined using a dual approach. Patients admitted during the first pandemic wave (March to September 2020) were categorized as having the ancestral (Wild type) strain based on the absolute epidemiological predominance in Spain at that time.

For subsequent waves, variant identification was performed through whole-ge-nome sequencing (WGS), or targeted genomic quantification (RT-PCR for specific mutations) conducted by the hospital’s Microbiology Department.

Vaccination status was retrieved from the regional health registry. Patients were classified according to the number of doses received prior to ICU admission: Un-vaccinated (0 doses), Partially Vaccinated (1 dose), Fully Vaccinated (2 doses)

#### 2.3.3 Clinical Severity and Organ Failure

Severity at admission was quantified using the Acute Physiology and Chronic Health Evaluation II (APACHE II) score, prospectively recorded in clinical practice, and SAPS II score, calculated retrospectively for the purposes of this study.

Organ dysfunction was assessed using the SOFA score, also calculated retro-spectively, at three intervals: the 24 hours preceding ICU admission (pre-ICU), at admission (first 24h), and at 72 hours.

Multiorgan failure was defined as the simultaneous acute dysfunction of two or more organ systems, characterized by a score of ≥2 points in each corresponding domain of the SOFA score.

#### 2.3.4 Clinical Management and Complications

Use of respiratory support like high flow nasal cannula (HFNC), non-invasive mechanical ventilation (NIMV) and invasive mechanical ventilation (IMV), prone positioning, neuromuscular blockade, and tracheostomy. Requirement of vasopres-sors, inhaled nitric oxide (iNO) and renal replacement therapy (RRT)

We also recorded major complications, such as pulmonary embolism and secondary infections including both bacterial or fungal etiology.

#### 2.3.5 Pharmacological Treatments

Administration of corticosteroids, tocilizumab, anakinra, and remdesivir. The drugs were administered according to the clinical protocols at the center during the study period, following the recommendations provided by the Spanish Agency of Medicines and Medical Devices (AEMPS).

### 2.4 Outcome Measures

The primary endpoint was all-cause hospital mortality, defined as death from any cause occurring between ICU admission and hospital discharge. Secondary out-comes were defined as follows:

- **Organ Dysfunction:** Incidence and severity of Multiorgan Failure (MOF), defined by SOFA criteria.
- **Duration of Organ Support:** Total ICU length of stay (LOS) and duration of organ support (e.g., days of invasive mechanical ventilation).
- **Clinical Complications:** Development of secondary bacterial or fungal infections and thromboembolic events (pulmonary embolism or deep vein thrombosis) during the hospital stay.

### 2.5 Statistical Analysis

A complete case analysis was performed. Given the very low rate of missing data (<1% of the cohort; 2 patients lacked MOF assessment at 24 hours, both of whom were non-survivors, presumably due to early death before evaluation), multiple imputation was not deemed necessary. The final regression model included 343 patients with complete data on all predictor variables.

Continuous variables are presented as median and interquartile range (IQR) or mean ± standard deviation (SD), as appropriate, based on their distribution. Categorical variables are expressed as counts and percentages.

Comparisons between survivors and non-survivors were performed using the Mann–Whitney U test or Student’s t-test for continuous variables, and the chi-square test or Fisher’s exact test for categorical variables, as appropriate.

A multivariable logistic regression model was constructed to identify independent predictors of hospital mortality. Variables were selected based on clinical relevance and statistical significance in univariate analysis (p < 0.10).

Severity scores and organ dysfunction variables were initially evaluated for multicollinearity; however, all were retained in the final model as they provided complementary prognostic information according to the AIC selection.

The linearity assumption between continuous predictors (SAPS II, age) and the logit of the outcome was assessed using the Box-Tidwell test. No significant departures from linearity were observed (p>0.05 for both variables), confirming that the inclusion of these variables in their original continuous form was appropriate.

A backward stepwise selection procedure based on the Akaike Information Criterion (AIC) was used to derive the final model.

Several measures were adopted to minimize potential sources of bias. Patients transferred between centers were excluded to ensure a complete clinical trajectory within our institution (see Patient Selection); the potential selection bias this entails is discussed in the Limitations.

To minimize confounding by indication, variables administered based on specific clinical severity criteria rather than randomization—such as tocilizumab—were evaluated separately and excluded from the final model when their inclusion did not reflect an independent causal association with the outcome (see Discussion).

Variable selection via AIC-based backward stepwise regression further reduced the risk of overfitting and spurious associations.

Sample size adequacy was evaluated using the Events Per Variable (EPV) criterion for multivariable prediction models (EPV ≥ 10) [18]. With 69 in-hospital deaths and 6 predictors in the final model (age, sex, COPD, hypertension, SAPS II, and MOF at 24 hours), the cohort yielded an EPV of 11.5, exceeding this threshold and supporting adequate power for stable coefficient estimation.

Model discrimination was evaluated using the A**r**ea Under the Receiver Operating Characteristic curve (AUC-ROC with corresponding 95% confidence intervals). The optimal cutoff point was determined using Youden’s J statistic.

The AUCs of nested models were compared with DeLong’s test, and the contribution of MOF at 24 h was also assessed with a likelihood ratio test. Calibration was evaluated with the Hosmer–Lemeshow test and a calibration plot.

All statistical analyses were performed using Python (v3.14.2), with the stats-models library (v0.14.6) for multivariable logistic regression via maximum likeli-hood estimation, and scikit-learn (v1.8.0) for ROC curve analysis. A two-tailed p-value < 0.05 was considered statistically significant.

### 2.6 Ethical Considerations

The study protocol was approved by the Research Ethics Committee with Medicinal Products (CEIm) of Hospital Universitario Príncipe de Asturias at its meeting of 19 March 2024 (letter dated 16 April 2024; Ref: CEIm OE 15/2024) and conducted according to the Declaration of Helsinki and its subsequent amendments (the latest in 2024), and in accordance with Good Clinical Practice (GCP) standards. Given the retrospective and anonymized nature of the data, the Committee accepted the waiver of informed consent.

## 3. RESULTS

### 3.1 Baseline characteristics

During the study period, a total of 469 patients were assessed for eligibility. Of these, 124 patients were excluded to ensure a cohort with a complete clinical trajectory and primary COVID-19 pathology.

Specifically, 94 patients were transferred to other hospitals due to ICU overca-pacity during pandemic peaks, 13 were excluded as they were transferred from other centers after a significant portion of their ICU stay had already elapsed, and 17 patients were excluded because SARS-CoV-2 infection was a concomitant finding rather than the primary cause for ICU admission.

Finally, a total of 345 critical COVID-19 patients admitted to the ICU were in-cluded in the study. The median age of the cohort was 62.0 years (IQR: 54.0– 69.0), with a predominance of male patients (64.3%). The overall in-hospital mortality rate was 20.6% (n=71). Obesity (62.3%) and hypertension (52.8%) were the most prevalent comorbidities in the overall cohort, followed by diabetes (23.2%).

Upon ICU admission, non-survivors were significantly older than survivors (66.0 [IQR 60.0–71.0] vs. 60.0 [IQR 52.0–68.0] years, respectively; p < 0.001) and pre-sented higher baseline severity scores, SAPS II (39.0 [IQR 33.0–46.0] vs. 31.0 [IQR 27.0–35.0]; p < 0.001) and APACHE II 15.0 [IQR 11.5–19.0 vs 11.0 [IQR 8.0–14.0]; p < 0.001).

Regarding baseline comorbidities, the Charlson Comorbidity Index score was higher in fatal cases (3.0 [IQR 2.0–4.0] vs. 2.0 [IQR 1.0–3.0]; p < 0.001), with COPD significantly more prevalent among non-survivors (23.9% vs. 13.9%; p = 0.046) (Table 1).

**Table 1.** Baseline demographic, clinical, and laboratory characteristics of the study cohort. Data are presented as median (IQR) for continuous variables and as number (%) for categorical variables. BMI: Body Mass Index; SAPS II: Simplified Acute Physiology Score II; APACHE II: Acute Physiology and Chronic Health Evaluation II; COPD: Chronic Obstructive Pulmonary Disease. P-values were calculated using the Mann-Whitney U test or Chi-square test, as appropriate.

| Variable | Total Cohort (N=345) | Survivors (N=274) | Non survivors (n=71) | p-value |
|---|---|---|---|---|
| **Demographics and Baseline Severity** |  |  |  |  |
| Age years Median IQR) | 62.0 (54.0–69.0) | 60.0 (52.0–68.0) | 66.0 (60.0–71.0) | **< 0.001** |
| Female Sex n (%) | 123 (35.7%) | 104 (38.0%) | 19 (26.8%) | 0.0952 |
| BMI, kg/m²<br>Median (IQR) | 30.0 (27.3–35.5) | 30.0 (27.4–35.5) | 30.0 (27.0–35.9) | 0.9696 |
| Charlson<br>Median (IQR) | 2.0 (1.0–4.0) | 2.0 (1.0–3.0) | 3.0 (2.0–4.0) | **< 0.001** |
| SAPSII 24 h ICU stay Median (IQR) | 32.0 (28.0–37.0) | 31.0 (27.0–35.0) | 39.0 (33.0–46.0) | **< 0.001** |
| APACHE II 24 h ICU stay Median (IQR) | 12.0 (9.0–15.0) | 11.0 (8.0–14.0) | 15.0 (11.5–19.0) | **< 0.001** |
| **Main Comorbidities, N (%)** |  |  |  |  |
| Obesity | 215 (62.3%) | 169 (61.7%) | 46 (64.8%) | 0.6815 |
| Hypertension | 182 (52.8%) | 138 (50.4%) | 44 (62.0%) | 0.0848 |
| Diabetes | 80 (23.2%) | 64 (23.4%) | 16 (22.5%) | 1.0000 |
| COPD | 55 (15.9%) | 38 (13.9%) | 17 (23.9%) | **0.0459** |
| CVD | 32 (9.3%) | 24 (8.8%) | 8 (11.3%) | 0.4964 |
| CKD | 21 (6.1%) | 17 (6.2%) | 4 (5.6%) | 1.0000 |
| Solid Cancer | 8 (2.3%) | 5 (1.8%) | 3 (4.2%) | 0.2141 |
| Hematologic Cancer | 15 (4.3%) | 11 (4.0%) | 4 (5.6%) | 0.5212 |
| HIV | 3 (0.9%) | 1 (0.4%) | 2 (2.8%) | 0.1088 |
| **Laboratory Findings at Admission, Median (IQR)** |  |  |  |  |
| Lymphocytes (×10³/µL) | 0.85 (0.63–1.11) | 0.84 (0.64–1.1) | 0.91 (0.61–1.23) | 0.6775 |
| Platelets (×10³/µL) | 184.0 (138.0–239.0) | 185.0 (146.0–231.0) | 179.0 (126.0–269.0) | 0.8706 |
| CRP (mg/L) | 102.4 (52.4–173.2) | 98.2 (52.1–172.3) | 125.0 (53.6–192.5) | 0.3140 |
| D-Dimer (mg/L) | 0.9 (0.6–1.6) | 0.9 (0.6–1.5) | 1.0 (0.7–3.4) | **0.0358** |
| Creatinine (mg/dL) | 0.9 (0.8–1.2) | 0.9 (0.8–1.2) | 1.0 (0.8–1.2) | 0.8431 |
| Ferritin (ng/mL) | 681.5 (360.8–1278.5) | 698.5 (391.0–1298.2) | 609.0 (250.5–1082.8) | 0.3732 |
| Bilirubin (mg/dL) | 0.5 (0.4–0.7) | 0.5 (0.4–0.7) | 0.5 (0.4–0.8) | 0.3732 |
| LDH (U/L) | 391.0 (300.5–509.5) | 386.0 (301.5–501.5) | 439.0 (298.5–534.5) | 0.3344 |
| Interleukin-6 (pg/mL) (n=188) | 60.2 (18.3–142.9) | 57.0 (18.1–133.5) | 123.7 (37.6–302.4) | **0.0341** |

At admission, laboratory analysis showed that non-survivors presented a worse systemic inflammatory profile, with elevated Interleukin-6 (IL-6) levels (123.7 [IQR 37.6–302.4] vs. 57.0 [IQR 18.1–133.5] pg/mL; p = 0.0341) (Table 1). Neither

SARS-CoV-2 variant (p = 0.372) nor vaccination status (p = 0.198) differed significantly between groups (Supplementary Table 1).

### 3.2 Clinical Course, ICU Management, and Survival

During the ICU stay, the development of acute MOF was a major factor associated with mortality. Pre-admission MOF was already more frequent among non-survivors (35.2% vs. 20.4%; p = 0.012), but this difference widened within the first 24 hours of ICU admission. Specifically, MOF at 24 hours was documented in 88.4% of non-survivors versus 61.7% of survivors (p < 0.001).

In terms of organ support, mechanical ventilation (IMV) was required in 95.8% of non-survivors, compared to 80.7% of survivors (p = 0.001). Also, non-survivors needed more use of vasopressors (93.0% vs. 77.0%; p = 0.002) and continuous renal replacement therapy (CRRT) (26.8% vs. 2.2%; p < 0.001). On the other hand, the use of high-flow nasal cannula (HFNC) was more frequent among survivors (57.3 % vs. 21.1%; p < 0.001).

Regarding complications, secondary fungal infections were significantly more prevalent in the non-survivor group (p = 0.002), whereas the incidence of pulmo-nary embolism and deep vein thrombosis did not differ significantly between groups (Supplementary Table 2).

**Table 2.**
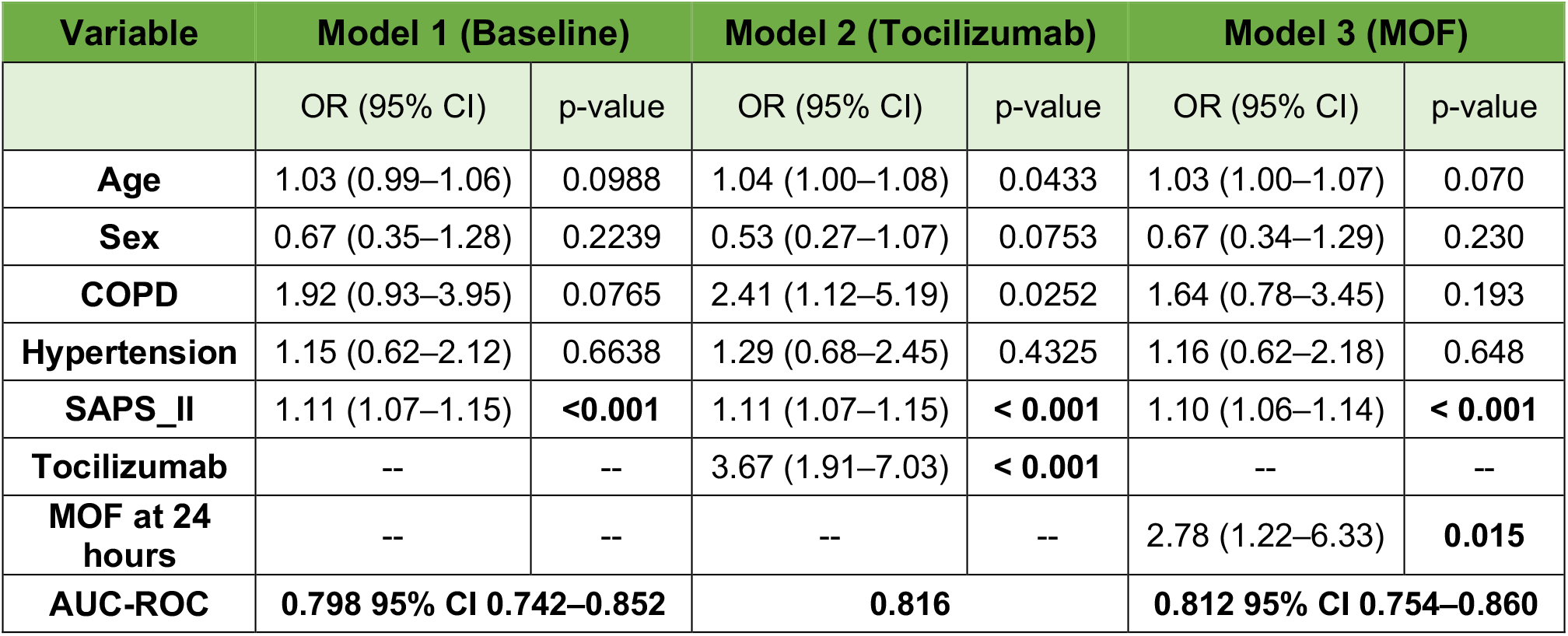
Multivariable logistic regression models for predictors of in-hospital mortality. OR: Odds Ratio; CI: Confidence Interval; AUC-ROC: Area Under the Receiver Operating Characteristic Curve. **Note:** The multivariable analysis included 343 patients, as two cases were excluded due to missing data.

### 3.3 Development of mortality model

To identify independent predictors of in-hospital mortality, a two-step modeling approach was used. An initial univariate screening was performed, followed by a multivariable logistic regression approach optimized by the Akaike Information Criterion (AIC).

To determine the best predictive model, three approaches were compared (Table 2). The baseline clinical model, based on patient demographics, comorbidities, and initial severity (age, sex, COPD, hypertension, and SAPS II), showed an acceptable predictive accuracy with an area under the curve (AUC) of 0.798 (95% CI 0.742–0.852).

Although adding Tocilizumab administration to the initial model improved its predictive capacity (AUC = 0.816), this variable was excluded from the final multivariable analysis to avoid confounding by indication bias.

Finally, the final model was built with baseline physiological severity and early dynamic clinical evolution, integrating the development of MOF at 24 h of admission (Table 2). This model had an AUC of 0.812 (95% CI 0.754–0.860) (Figure 2A), which was not statistically different from the baseline model by DeLong’s test (ΔAUC = +0.014, p = 0.225); however, the likelihood ratio test showed that adding MOF at 24 h significantly improved model fit (χ^2^ = 6.79, p = 0.009). In the adjusted analysis, early MOF development was a major determinant of mortality, almost tripling the odds of in-hospital death (OR =2.78 [1.22-6.33] p = 0.015). (Table 2 and Figure 2B).

**Figure 1.**
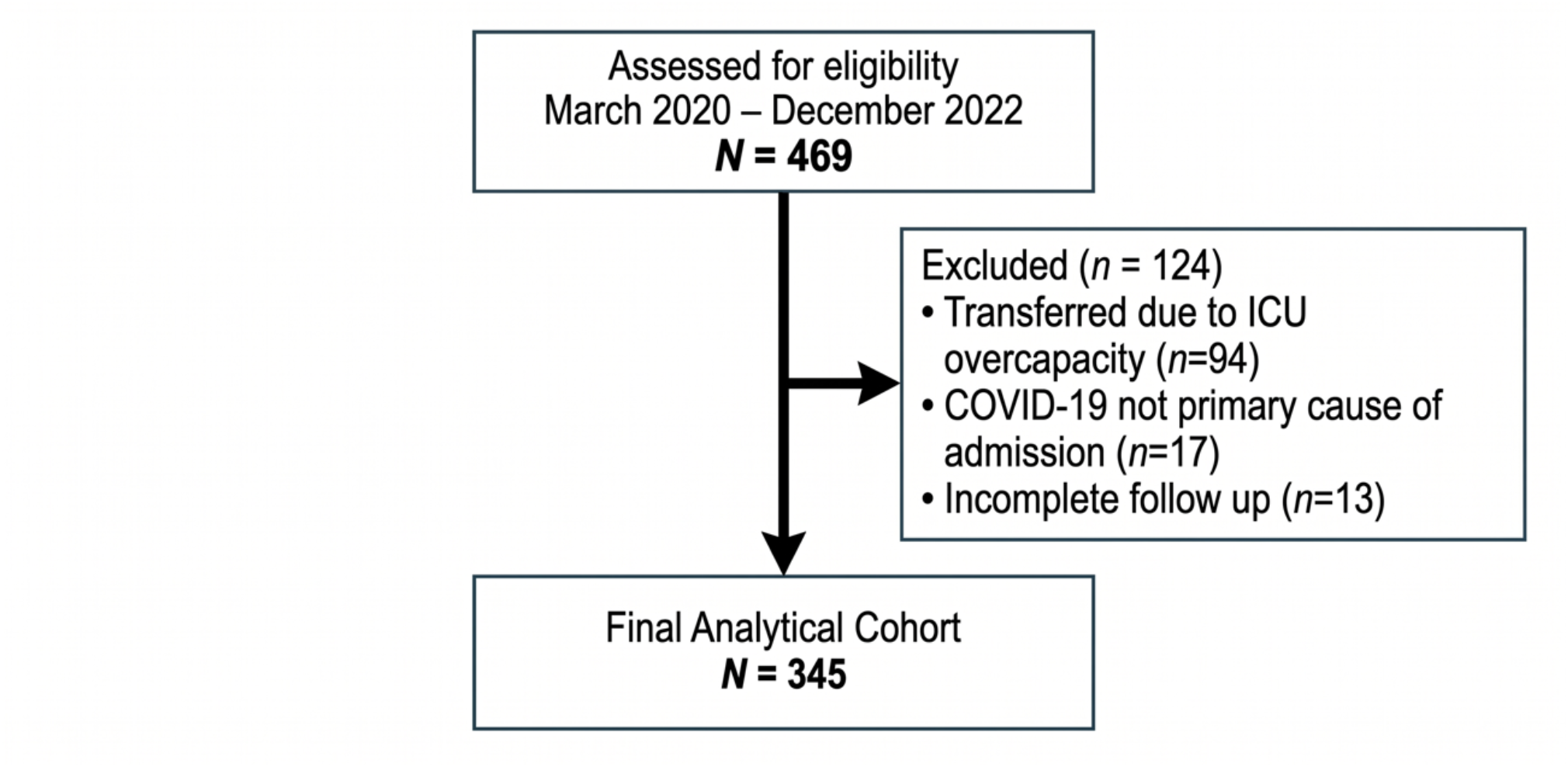
STROBE Flow Diagram of Patient Selection

**Figure 2.**
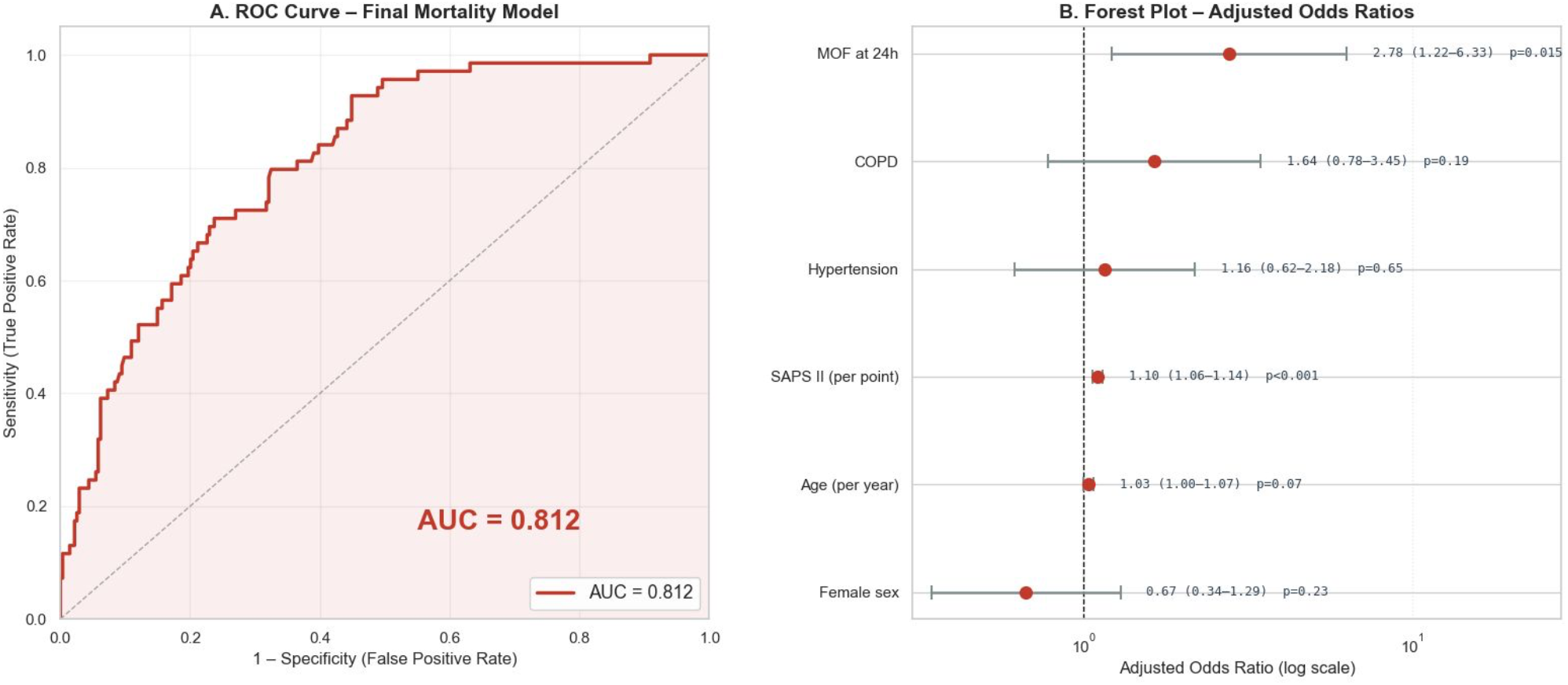
Predictive value and multivariable risk factors for in-hospital mortality. (2A) AUC-ROC of the definitive model (AUC = 0.812). (2B) Forest plot representing the multivariable adjusted Odds Ratios (OR) and 95% confidence intervals for the independent predictors of mortality.

Using Youden’s index, the optimal probability threshold of the final model was 0.116, yielding a sensitivity of 92.8% and a specificity of 55.1%. The Hosmer– Lemeshow test showed no significant departure from perfect calibration (χ^2^ = 13.50, p = 0.096), and the calibration plot showed adequate agreement between predicted and observed mortality.

The individual and adjusted contribution of each variable to hospital mortality is summarized in the Forest Plot (**Figure 2B**).

## 4. Discussion

In this study, we developed a practical clinical model to predict hospital mortality in critically ill COVID-19 patients. Our findings indicate that combining clinical patient severity with the development of MOF in the first 24 hours of ICU admission provides good discriminative ability.

Our study population showed a clinical profile similar to that observed in other major series, with older age and a greater comorbidity burden among non-survivors compared with survivors [5,7,19]. Furthermore, as expected, non-survivors required more organ support and prolonged ICU stays. Regarding mortality, while previous meta-analyses have reported pooled in-hospital mortality rates of approximately 32–34%-with individual cohorts varying widely between 4.5% and 69.5% [1,2] - the overall mortality in our cohort was 20.6%. These findings allow direct comparison of our results with other critically ill populations.

When evaluating the variables in our predictive models, SAPS II emerged as a strong and consistent predictor of mortality across all three models consistent with its established role as a severity score in critically ill patients [9,20,21]. Although models were adjusted by sex and age, they were not independently associated with mortality in our cohort. Age is already included in the SAPS II calculation, meaning both variables may be collinear. Likewise, hypertension was retained in all three models despite not reaching statistical significance, given its established association with mortality [22].

During the development of our predictive tool, we explored incorporating tocilizumab use into the model (Model 2) due to its strong association with mortality in preliminary analyses. While its inclusion produced a marginal increase in discrim-inative performance (AUC), this finding, contradicts the evidence supporting its therapeutic benefit in severe COVID-19 [23,24].

Tocilizumab in our institution was administered exclusively on the general ward prior to ICU admission, following a hospital protocol based on Spanish Society of Infectious Diseases and Clinical Microbiology (SEIMC) recommendations (Supplementary Appendix 1) [25]. This targeted administration suggests confounding by indication; patients who received tocilizumab already met criteria for severe hyperinflammatory disease before ICU admission, probably making it a marker of extreme severity rather than a causal contributor to death. This indication bias has been proposed before in the case of tocilizumab: a large multicenter cohort of 422 critically ill COVID-19 patients from Andalusian ICUs found no independent association between tocilizumab use and improved survival [14]. Additionally, because the drug was administered before ICU admission, its exact timing could not be determined. Given this bias and the evolution of treatment protocols, we excluded tocilizumab from the final model.

In the final model (Model 3), we replaced tocilizumab with MOF at 24 hours of ICU admission. Early MOF proved to be an independent predictor of mortality without compromising the performance of the model. However, COPD’s association with mortality weakened once MOF was introduced, suggesting its effect may be partly mediated by early organ dysfunction. Chronic respiratory disease depletes patient’s physiological reserve, facilitating a much faster progression to systemic organ failure [26]; however, this hypothesis would require further studies.

Taken together, these findings support our central hypothesis: mortality risk is best defined by a combination of comorbidities --such as hypertension, male sex and COPD--, severity at admission and early organ dysfunction. Traditional scores like SAPS II and SOFA provide only a static picture and may fail to fully capture the dynamic course of systemic COVID-19 [27]. Relying solely on base-line scores may not be enough; it seems that incorporating early dynamic evaluation is crucial to capture the true risk of mortality [13].

Our results support the idea of critical COVID-19 as a systemic syndrome and not only as a respiratory disease (as believed initially), causing endothelial injury and a dysregulated inflammatory response that can rapidly lead to multiorgan failure [11,12]. Assessing organ dysfunction at 24 hours identifies patients who fail to stabilize despite initial supportive treatment, offering a dynamic complement to static admission scores.

Several models have been proposed to predict mortality in COVID-19. Among clinical severity scores, Moisa et al. developed the COVID-SOFA score for 28-day mortality in a cohort of 425 ICU patients, combining age, neutrophil-to-lym-phocyte ratio, and SOFA score into a single equation, improving on the SOFA score alone [15]. Our model achieves comparable performance using simpler variables at a single 24-hour assessment.

Other authors have explored machine-learning approaches. Aznar-Gimeno et al. used XGBoost in a large hospitalized COVID-19 cohort (n=3623), reducing an initial pool of 165 candidate variables—spanning demographics, comorbidities, chronic medication, vital signs, and laboratory data—to a final set of 20 predictors selected by variable importance [28]. A two-center study from the University of Washington and Tongji Hospital with a similar approach, included variables collected within the first 24 hours of admission, including age, white blood cell count, and platelet count [29]. While these models can be similar or exceed our discrim-inative performance, they typically require larger variable sets, selection procedures, or complex algorithms that make these variables difficult to interpret together, limiting bedside interpretability.

Beyond its statistical performance, this model could, once validated, serve as a practical aid for early risk stratification, as it includes simple variables routinely calculated in the ICU without adding extra workload, informing, but never replacing, clinical judgment on escalation of care or, when appropriate, on the adequacy of therapeutic effort. This is especially relevant today, in contemporary ICUs that now admit comorbid, fragile patients in which deciding when to stop treatment is often particularly difficult.

This study has several strengths, including a well-characterized, homogeneous cohort, the early assessment of organ dysfunction, which may reduce survivor-ship bias, and its direct clinical applicability.

However, certain limitations should be acknowledged. First, the retrospective, single-center design and the absence of external validation limit the generalizability of our findings, and the cutoff point was derived in the same cohort used to develop the model.

Second, 94 patients (20% of those initially assessed) were transferred to other hospitals and therefore excluded. These transfers were driven mainly by ICU overcapacity during pandemic peaks and only exceptionally by clinical severity. Although this suggests that transfer was largely unrelated to prognosis, it still constitutes a non-random selection step: if clinical stability influenced which patients were transferred, the case-mix of the final cohort (N=345) may differ from the full population of critically ill COVID-19 patients at this center, which should be considered when interpreting the observed mortality (20.6%).

Finally, the potential confounding effect of sedation and vasopressor use on organ dysfunction and the evolving treatment protocols during the pandemic, which may have changed baseline mortality risk as the standard of care progressed, should also be taken into account.

## 5. Conclusion

In summary, combining baseline clinical severity with the early development of multiorgan failure provides a simple risk stratification tool that, pending external validation, could assist in daily decision-making in the critically ill COVID-19 population.

## Data Availability

The data supporting this study contain individual-level clinical information from patients and are not publicly available due to privacy restrictions. Anonymized data are available from the corresponding author upon reasonable request, subject to approval by the Research Ethics Committee with Medicinal Products (CEIm) of Hospital Universitario Principe de Asturias.

## Data Availability Statement

The data supporting this study contain individual-level clinical information from patients and are not publicly available due to privacy restrictions. Anonymized data are available from the corresponding author upon reasonable request, subject to approval by the Research Ethics Committee with Medicinal Products (CEIm) of Hospital Universitario Príncipe de Asturias.

## Declaration of generative AI and AI-assisted technologies in the manuscript preparation process

During the preparation of this work the author(s) used Gemini and Claude to improve English grammar and assist with statistical coding and image preparation. After using this tool, the author reviewed and edited the content as needed and take full responsibility for the content of the published article.

