## Supplementary Material for "Baseline Severity and Early Multiorgan Failure to Predict In-Hospital Mortality in Critically Ill COVID-19 Patients: A Retrospective Cohort Study"

### Supplementary Table S1 SARS-CoV-2 Variant Distribution and Baseline Vaccination Status

| Variable | Total Cohort (N=345) | Survivors (N=274) | Non survivors (n=71) | p-value |
|---|---|---|---|---|
| **COVID-19 Characteristics, n (%)** | | | | p=0.3722 |
| Ancestral variant (Wild type) | 215 (62.3%) | 161 (58.8%) | 54 (76.1%) | |
| Delta | 18 (5.2%) | 16 (5.8%) | 2 (2.8%) | |
| B117 | 32 (9.3%) | 27 (9.9%) | 5 (7.0%) | |
| Other | 3 (0.9%) | 2 (0.7%) | 1 (1.4%) | |
| Unknown | 77 (22.3%) | 68 (24.8%) | 9 (12.7%) | |
| **Vaccination Status, n (%)** | | | | p=0.1985 |
| Unvaccinated (0 doses) | 309 (89.6%) | 247 (90.1%) | 62 (87.3%) | |
| Partially vaccinated (1 dose) | 6 (1.7%) | 3 (1.1%) | 3 (4.2%) | |
| Vaccinated (2 doses) | 30 (8.7%) | 24 (8.8%) | 6 (8.5%) | |

Categorical variables are expressed as frequencies and percentages (n, %)

**Supplementary Table S2. Clinical Course, Organ Support, Complications and Treatments in the ICU.**

| Variable | Total Cohort (N=345) | Survivors (N=274) | Non survivors (n=71) | p-value |
|---|---|---|---|---|
| **Organ Dysfunction Dynamics** | | | | |
| MOF pre-ICU n (%) | 81(23.5%) | 56 (20.4%) | 25 (35.2%) | **0.0118** |
| SOFA pre-ICU Median (IQR) | 3.0 (2.0–4.0) | 3.0 (2.0–3.8) | 3.0 (2.0–4.0) | 0.2276 |
| MOF at 24h n (%) | 230/343 (67.1%) | 169/274 (61.7%) | 61/69 (88.4%) | **< 0.001** |
| SOFA 24h Median (IQR) | 5.0 (3.0–7.0) | 5.0 (3.0–6.0) | 7.0 (5.0–8.0) | **< 0.001** |
| **Organ Support and Interventions, n (%)** | | | | |
| IMV | 289 (83.8) | 221(80.7%) | 68 (95.8%) | **0.0010** |
| Tracheostomy | 129 (37.4) | 104 (38.0%) | 25 (35.2%) | 0.7833 |
| HFNC | 172 (49.9) | 157 (57.3%) | 15 (21.1%) | **< 0.001** |
| Prone positioning | 184 (53.3) | 125 (45.6%) | 59 (83.1%) | **< 0.001** |
| Neuromuscular blockade | 237 (68.7) | 173 (63.1%) | 64 (90.1%) | **< 0.001** |
| Vasopressors | 277 (80.3) | 211 (77.0%) | 66 (93.0%) | **0.0022** |
| CRRT | 25 (7.2) | 6 (2.2%) | 19 (26.8%) | **< 0.001** |
| **Clinical Complications, n (%)** | | | | |
| Secondary bacterial infection | 202 (58.6) | 153 (55.8%) | 49 (69.0%) | 0.0580 |
| Secondary fungal infection | 7 (2.0) | 2 (0.7%) | 5 (7.0%) | **0.0016** |
| PE | 48 (13.9) | 38 (13.9%) | 10 (14.1%) | 1.0000 |
| DVT | 16 (4.6) | 11 (4.0%) | 5 (7.0%) | 0.3381 |
| **COVID-19 Specific Therapies, n (%)** | | | | |
| Corticosteroids (any dose) | 317 (91.9) | 257 (93.8%) | 60 (84.5%) | **0.0248** |
| Corticosteroid pulse therapy | 52 (15.1) | 31 (11.3%) | 21 (30.0%) | **0.0003** |
| Tocilizumab | 154 (44.6) | 109 (39.8%) | 45 (63.4%) | **0.0005** |
| Remdesivir | 40 (11.6) | 33 (12.0%) | 7 (9.9%) | 0.6832 |
| **Outcomes** | | | | |
| Duration of IMV, days Median (IQR) | 11.0 (5.0–22.0) | 10.0 (5.0–22.0) | 14.0 (8.0–21.5) | 0.1165 |
| ICU length of stay, days Median (IQR) | 14.0 (7.0–31.0) | 14.0 (7.0–31.0) | 15.0 (6.0–27.0) | 0.7713 |

Data are presented as median (interquartile range (IQR)) or n (%). p-values indicate univariable comparisons between survivors and non-survivors. Abbreviations: CRRT, Continuous Renal Replacement Therapy; DVT, Deep Vein Thrombosis; HFNC, High-Flow Nasal Cannula; IMV, Invasive Mechanical Ventilation; MOF, Multiorgan Failure; PE, Pulmonary Embolism; SOFA, Sequential Organ Failure Assessment.

### Supplementary Table S3. Observed mortality and log-odds across quartiles of continuous predictors, and formal linearity testing (Box-Tidwell test).

| Variable | Quartile | N | Deaths, n (%) | Empirical Logit | Box-Tidwell p-value |
|---|---|---|---|---|---|
| SAPS II | Q1 | 95 | 4 (4.2%) | -3.125 | 0.704 |
| | Q2 | 87 | 13 (14.9%) | -1.739 | |
| | Q3 | 78 | 11 (14.1%) | -1.807 | |
| | Q4 | 85 | 43 (50.6%) | +0.024 | |
| Age (years) | Q1 | 100 | 7 (7.0%) | -2.587 | 0.436 |
| | Q2 | 86 | 18 (20.9%) | -1.329 | |
| | Q3 | 84 | 22 (26.2%) | -1.036 | |
| | Q4 | 75 | 24 (32.0%) | -0.754 | |

Empirical logit = $\ln[p/(1-p)]$. Linearity was assessed by the Box-Tidwell test; $p > 0.05$ indicates no deviation from linearity.

### Supplementary Figure S1. Model calibration and predicted probability distributions.

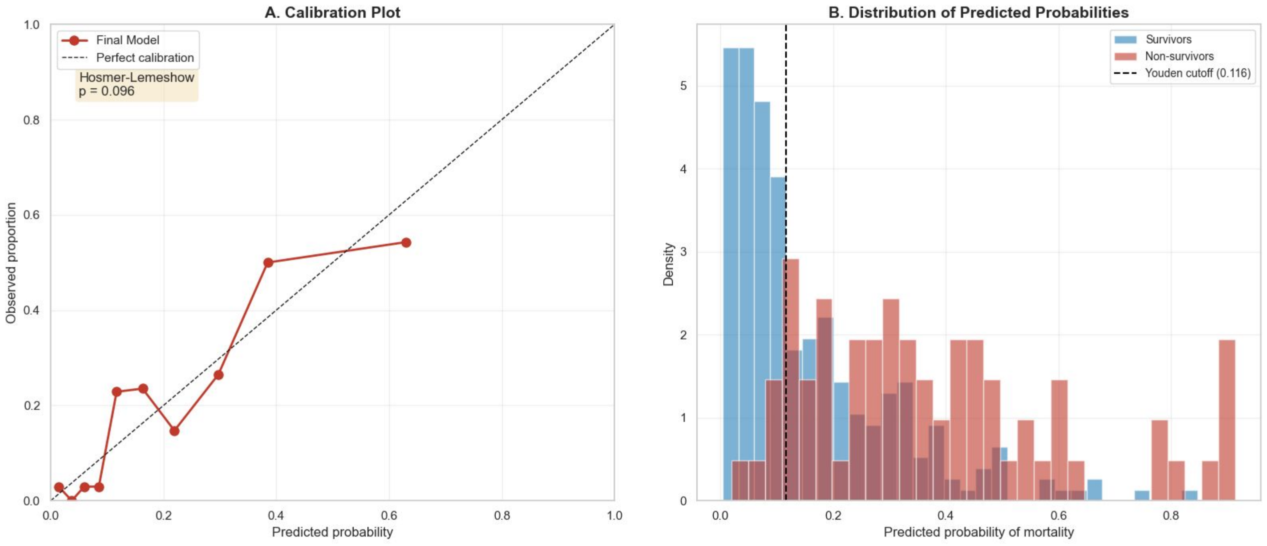


**Calibration plot and predicted probabilities distribution.** Left: Calibration curve of the final model (Hosmer–Lemeshow p = 0.096); the dashed diagonal represents ideal calibration. Right: Density distribution of predicted mortality probabilities for survivors and non-survivors. The vertical dashed line marks the optimal Youden cutoff (0.116).

### Supplementary Appendix S1. Protocolized Criteria for Tocilizumab Administration

During the study period, the administration of tocilizumab in our institution followed a standardized clinical protocol based on objective indicators of hyperinflammatory state prior to ICU admission. Patients were considered eligible for treatment if they met the following criteria:

Eligible patients required RT-PCR-confirmed infection with radiological infiltrates and clinical worsening (oxygen saturation ≤94%) and/or biomarker evidence of hyperinflammation.

- IL-6 >40 pg./mL
- Ferritin >500 ng/mL
- D-dimer >1.5 µg/mL
- Troponin >0.12 ng/mL
- CRP >150 mg/L
- LDH >245 U/L
- Lymphocyte count <800/µL
- Neutrophil-to-lymphocyte ratio >3.13).
